# A Semi-Supervised Contrastive Learning Approach to Alzheimer’s Disease Diagnostics using Convolutional Autoencoders

**DOI:** 10.1101/2022.12.27.22283984

**Authors:** Edward Jung, Anshul Kashyap, Brandon Hsu, Mason Moreland, Chanon Chantaduly, Peter D. Chang

**Author notes:** **Subject Overlap:** None.

## Abstract

**PURPOSE:** Alzheimer’s Disease (AD) is a neurodegenerative disease that progressively deteriorates memory and cognitive abilities. PET 18F-AV45 (florbetapir) is a common imaging modality used to characterize the distribution of beta-amyloid deposits in the brain, however interpretation may be subjective and the misdiagnosis rate of AD ranges from 12-23%. Automated algorithms for PET 18F-AV45 interpretation including those derived from deep learning may facilitate more objective and accurate AD diagnosis.

**MATERIALS & METHODS:** A total of 1232 PET AV45 scans (207 - AD; 1025 - normal) were obtained from the Alzheimer’s Disease Neuroimaging Initiative (ADNI). A semi-supervised deep learning framework was developed to differentiate AD and normal patients. The framework consists of an autoencoder (AE), a contrastive learning loss, and a categorical classification head. A contrastive learning paradigm is used to improve the discriminative properties of latent feature vectors in multidimensional space.

**RESULTS:** Upon five-fold cross-validation, the best-performing semi-supervised contrastive model achieved validation accuracy of 82% to 86%. Secondary analysis included visualization of intermediate activations, classification report verification, and principal component analysis (PCA) of latent feature vectors. The training process yielded optimal converging losses for all three loss frameworks.

**CONCLUSION:** A deep learning model can accurately diagnose AD using PET 18F-AV45 scans. Such models require large amounts of labeled data during training. The use of a semi-supervised contrastive learning objective and AE regularizer helps to improve model performance, especially when dataset sizes are constrained. Latent representations extracted by the model are visually clustered strongly with the addition of a contrastive learning mechanism.

**Summary Statement:** A semi-supervised contrastive learning deep learning system optimizes latent feature vector representations and yields strong model classification performance for larger data distributions within the Alzheimer’s Disease diagnostics domain.

**Key Points:**

1. A common diagnostic procedure used by trained radiologists in the clinical setting is the visual analysis of PET 18F-AV45 neuroimaging scans to diagnose the different stages of Alzheimer’s Disease in a patient.
2. Contrastive learning is a strategy that allows for the optimization of latent feature representations in multidimensional space through the use of a loss function that maximizes the distance between feature vectors of different classes and minimizes the distance of feature vectors of the same class.
3. A semi-supervised contrastive learning approach can lead to improved performance and generalization of deep learning models optimized using small training datasets as encountered in Alzheimer’s Disease and other neurodegenerative conditions.

## INTRODUCTION

According to the Alzheimer’s Association, more than 6 million individuals were living with Alzheimer’s Disease (AD) in 2021 [1]. AD is misdiagnosed at a high rate of 12-23% [2] and is one of the top 10 leading causes of death in the United States [3]. In addition to memory problems, AD presents an array of symptoms that can seriously disrupt an individual’s life [3]. In the clinical environment, AD is commonly diagnosed through the analysis of neuroimaging scans by trained radiologists [4]. Specifically, PET AV45 (florbetapir), a neuroimaging modality commonly used to identify beta-amyloid clusters in a subject’s brain, is used by radiologists due to the correlations between beta-amyloid quantities & locations and the progression of AD [5]. However, subjective biases which are inherent in radiologist-based diagnostics cause issues with the generality of AD diagnostics [6].

Deep Learning (DL) is a subset of machine learning that focuses on utilizing artificial neural networks to generalize across large/varying data distributions [7]. In recent years, the use of DL neural networks within clinical diagnostics have yielded strong results in diagnostic accuracy and speed. Within the AD diagnostics domain, DL models have proven to successfully perform disease progression predictions. A study by Huang et al yielded consistent accuracies of 84.5% utilizing PET FDG (metabolism) scans for AD vs. CN classification [8-9]. PET AV45 scans can be used in the DL spectrum in training neural networks to identify specific features/biomarkers and automate the diagnosis task. DL approaches which have tackled this problem frequently utilize end-to-end 3D CNN architectures for feature extraction and classification subject only to cross-entropy classification loss or a combination of AEs and classifications heads [10]. These existing approaches commonly use single loss methodologies on small datasets and are subject to class imbalances (there are usually always significantly more CN than AD neuroimaging scans available), limiting the model’s ability to generalize and form effective decision boundaries [11-12].

This study proposes the use of a contrastive learning paradigm to accelerate extracted feature differentiation and to optimize the distances between latent feature vectors of differing and similar input samples [13-14]. We utilize a semi-supervised deep learning system consisting of an AE, a binary classification head, and a contrastive learning head. The AE is used to reduce the dimensionality of the inputted PET 18F-AV45 neuroimaging scan into a latent feature vector with extracted features that are key to the classification task at hand - a multi-loss training process is comparatively faster and retains high test accuracies [15]. The classification head performs binary predictions (Alzheimer’s Disease (AD):Control (CN)) on the extracted latent feature vectors. The contrastive learning head computes a distance metric between the latent feature vectors and creates gradients that correspond to the ideal distance between the feature vectors (maximum or minimum).

These optimal positions which are created due to the emphasized distance loss which propagates position-aware gradient computation can be visually discerned through PCA reduced 2D plots [16]. Further hyperparameter tuning to improve individual subsystem performances can yield optimal results. Specifically, we hypothesize that contrastive learning can contribute to high classification accuracies with the training process accelerated by the addition of AE reconstruction loss.

## MATERIALS & METHODS

### DATA & ANNOTATIONS

A total of 1232 PET 18F-AV45 samples were obtained from the open-source ADNI and clustered into two different collections [17]. There were 207 samples in the positive cohort (subjects with AD) and 1025 samples in the negative cohort (subjects classified as CN) and a 4:1 train:validation split was utilized during the training process. Due to the noticeable imbalance in the different class distributions, a custom data generator was created to maintain a 50:50 input distribution of AD and CN samples during the training and validation processes. The positive cohort was defined to be the subjects with Alzheimer’s Disease whereas the negative cohort was defined to be the subjects falling under the “Control” category (individuals without Alzheimer’s Disease). The data was contributed to the ADNI from PET Core (University of Michigan) group and was preprocessed prior to downloading the CN and AD collections on our local system [18]. PET 18F-AV45 samples were pre processed prior to downloading the AD and CN collections to our local system and the standardization techniques that were applied to each sample are shown in **table 1**.

**Table 1.** Preprocessing Techniques Employed Prior to Collection Download. This table shows all the preprocessing techniques used to standardize the AV45 samples prior to downloading the data on the system this study used.

|  | Process Name | Package Name | Program Name |
| --- | --- | --- | --- |
| **Step 1** | Smooth | Xida | cvt_ext_smooth |
| **Step 2** | Calculate Coregistration | Neurostat | mcoreg |
| **Step 3** | Apply Coregistration | Neurostat | coregimg |
| **Step 4** | Average Frames | Xida | ave_sep_frames |
| **Step 5** | AC-PC Orient Baseline | Neurostat | stereo |
| **Step 6** | Standardize to Baseline | Neurostat | coreg |
| **Step 7** | Average Frames | Xida | avg_sep_frames |
| **Step 8** | Intensity Normalization to Cerebellar<br>Gray Matter | Xida | ImageNormalize |
| **Step 9** | Variable Smooth | Xida | SmoothFile |

### LOCAL PREPROCESSING

The data was subsequently preprocessed to a shape of 96×128×128 with a Z-dimension slice thickness of 3mm with no additional padding and then normalized using Z-score normalization. All images are ensured to be axial. During the training process, stratified sampling was set to be 0.5 AD and 0.5 CN. Randomized shifting and scaling was also set to 20%. These steps were taken to further improve model training efficiency and to ensure that the dataset would not skew the training sequence.

### CORE ARCHITECTURE

Two types of experiments were performed with varying hyperparameters and different model architectures. In order to simplify the task at hand and isolate the different components to easily discern the performance metrics of the crucial/necessary system, the initial iteration of the experiment did not contain reconstruction loss. Reconstruction loss was added to the system once an optimal model with only contrastive and cross-entropy loss was found after a thorough hyperparameter sweep. Cosine similarity, euclidean distance, and a weighted combination of the two were all tested to determine the optimal difference metric for contrastive loss [19-20].

Cross-entropy loss and contrastive loss with cosine similarity as the difference metric were used in a dual loss training process to simultaneously train the classification head and improve latent space vector positioning. The AE architecture used during this experiment is presented in **figure 1**. Four encoder and four decoder blocks were used to produce latent space feature representations of the input sample and reconstruct the input sample from the feature vector. It is important to note that the computed weights did not take into account reconstruction loss (reconstruction loss was weighted 0 for the entire training process) for the first iteration of the experiment. Each encoder block contained a 3D convolution layer with a kernel size of (3, 3, 3) followed by a Leaky ReLU (alpha value of 0.3) activation layer. A batch normalization layer was used to improve the model’s ability to generalize on a larger input data distribution and to accelerate the training process as a whole by normalizing intermediary outputs. Due to the limited dataset size, an additional l2 regularization methodology was employed to prevent overfitting and ensure the model’s ability to generalize across a larger data distribution during inference [21].

**Figure 1.**
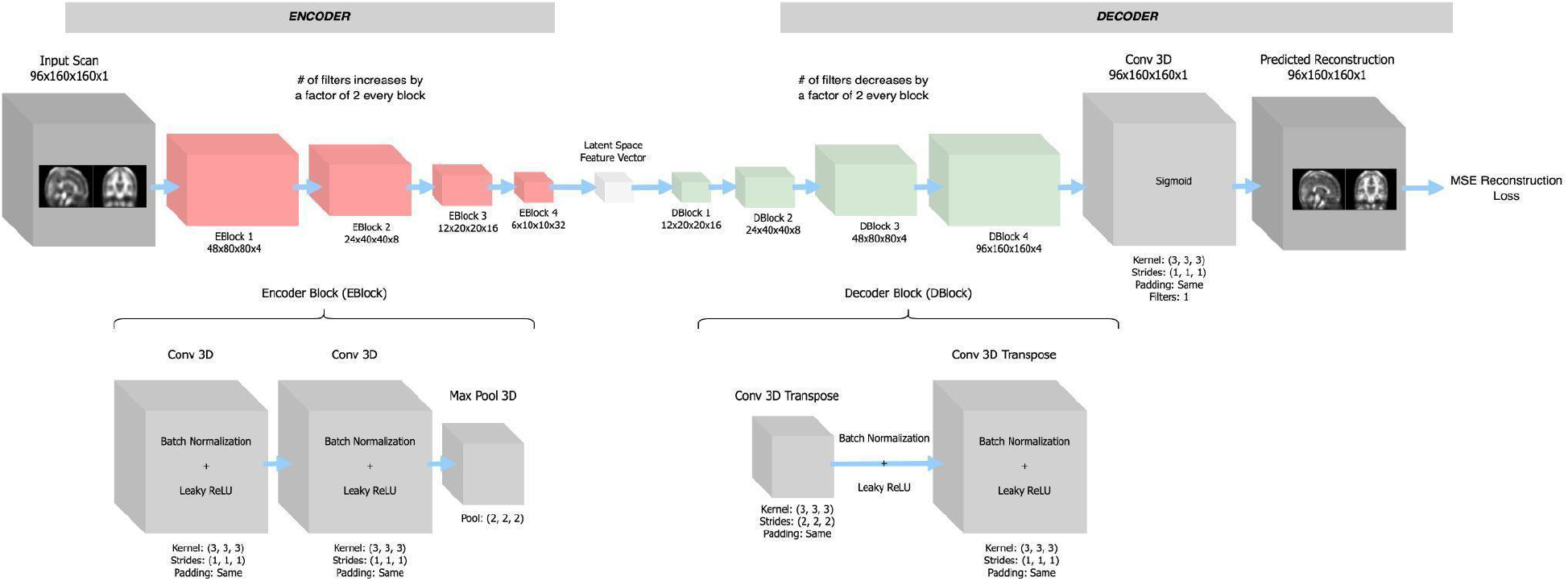
3D AE Architecture. Experiments with a limited training dataset distribution size tend to leverage the power of autoencoder reconstruction loss to accelerate the feature extraction process. The reconstruction task forces the model to learn meaningful low rank approximations in order to reconstruct the input sample. The model will learn important features for the classification task and the reconstruction task when coupled with classification loss. The figure above provides an overview of the AE architecture used in this study for latent feature extraction.

### ADDING RECONSTRUCTION

Once stable loss and accuracy values were achieved, reconstruction loss was added as a normalization technique and a catalyst to speed up the training process as a whole. The supervised contrative learning system architecture is shown in **figure 2**. The combination of reconstruction and contrastive loss in a triple loss system acted as a regularizer to improve the generality of the diagnostic model by swaying the model’s training process away from focusing solely on the classification problem to reduce the chances of overfitting.

**Figure 2.**
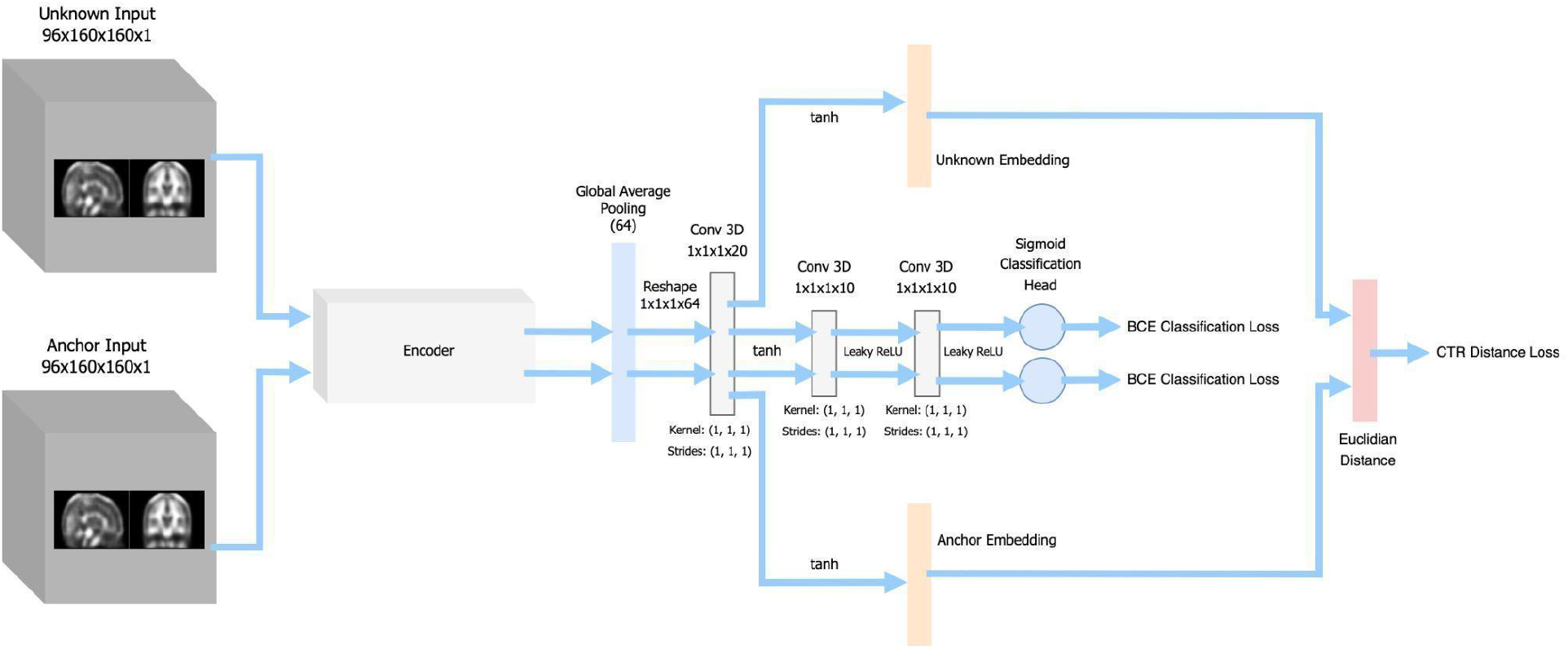
Contrastive Learning + Classification System Architecture. This figure highlights the data flow during the contrastive learning and classification training processes performed in this experiment. The extracted features from the anchor and unknown samples passed into the contrastive learning head where a distance metric (cosine similarity and Euclidean distance) was computed between elements of the feature vector in the same position. Binary cross-entropy loss was simultaneously computed for each of the predicted classes for the latent feature vectors.

### IMPLEMENTATION

Mean squared error (MSE) reconstruction loss was implemented to accelerate the AE’s training process and to extract latent features quickly and efficiently. To ensure that the extracted feature vectors were useful to the classification task at hand, and did not transform into a generic identity function, a triple loss training methodology was used. By effectively imposing a set of restrictions on the reconstruction problem, an accelerated pathway was created to minimize binary classification (AD:CN) log loss. Cross-entropy loss was also used when incorporating the contrastive learning paradigm. A distance metric was first computed by calculating the euclidean distance and/or cosine similarity between the extracted latent feature vectors. Since the individual values within the latent feature vectors were passed through a softmax function, generating normalized distance values between 0 and 1 respectively.

Cross-entropy contrastive loss was then computed using one minus the normalized distance being the predicted label and whether the binary classification labels were equal to each other or not being the true label. Loss weights were applied to modify the overall effect each of the three losses had on the model’s training process: [1.0, 0.5, 0.75] mapped to [binary classification loss, contrastive loss, and reconstruction loss]. The Adam method was used to optimize the model with a learning rate of 5E-6 and a decay rate of 0.03 [22]. To prevent a preference for the class with a larger number of samples (CN - 1025 samples), a custom generator was created to produce a 50:50 input sample distribution. Model weights were randomly initialized at the beginning of each training process using the Xavier distribution [23].

This study utilized a batch size of 8 paired scans (4 training examples total per iteration) and the training process was run with a step size of 50 for 4000 iterations (80 epochs).

The models were developed using Python Version 3.6 RRID:SCR_008394, TensorFlow [24] Version 2.1.0 RRID:SCR_0163, and Keras Version 1.0.8. All experiments were performed using a total of 8 NVIDIA Titan RTX 24 GB GPU cards. The average model training time for each experiment was 4 hours, while the time for single inference prediction was 0.113 seconds.

### STATISTICS

Binary cross-entropy log loss was used for both the classification and contrastive tasks. Binary log loss values calculated from the logits (capped between zero and one) produced by the classification head and the labels were used during the training process to improve the model’s classification performance. For the contrastive loss paradigm, the latent feature representation output of the encoder of the AE was first computed for both the unknown and anchor scans.

The distance between these two extracted vectors was subsequently derived with varying difference algorithms: euclidean distance, cosine similarity, and a weighted average of both. A sigmoid activation head was then used to cap the output value of the contrastive learning head between zero and one. Binary log loss values calculated from the logits produced by the contrastive head and the labels (zero: the classification labels are the same; one: the classification labels are different) were used during the training process to produce optimal latent space vector positions. Each prediction from the anchor and unknown AE is compared to the ground truth through binary log loss.

To measure the performances of this study’s models in-depth, a classification report was produced after each experiment which included the following metrics: weighted f1, regular f1, sensitivity, specificity, predictive positive value (PPV) and negative predictive value (NPV). In order to see how the model performed according to various logit threshold values, receiver operating characteristic and precision recall curve graphs were generated. To compare models, weighted f1, area under receiver operating characteristic (AUROC), and area under precision recall curve (AUPRC) statistics were used. Other analysis methods, such as utilizing the PCA dimensionality reduction technique to visualize latent feature vectors in 3D space, analyzing model extracted features, and reconstruction results, were crucial to figuring out if specific components of the system were functional.

## RESULTS

To evaluate the performance of the different training methodologies, a static model was trained with different sets of loss function weights and evaluated on a test set. Because it is possible that the performance of a specific training methodology may be influenced by the exact model hyperparameters used, 16 different model hyperparameter architectures were trained and evaluated for each of the loss functions. The space of models tested are shown in **table 2** and the aggregated results are shown in **table 3**. Identifying the proper combination of hyperparameters was just as important as optimizing specific hyper parameter values as the performance of the model directly correlated to the relationship between hyperparameters as well.

**Table 2.** Model Component Dimensions and Activation Function Details. This table provides information about the space of embedding and projection dimensions chosen for the models during this study. In total, this table represents 16 different hyperparameter configurations.

| Model Permutations | Cartesian Product | Description |
| --- | --- | --- |
| Projection Dimension | 256, 128, 64, 32 | layer after pooling |
| Embedding Dimension | 64, 32, 16, 8 | layer after projection |

**Table 3.** Single Classification Loss/Contrastive Learning Boosted Classification Statistics. This table shows the classification performance statistics on the three experimental models (no-ctr → no contrastive, ctr-euc → Euclidean distance, and ctr-cos → cosine similarity).

| Loss 1 | Loss 2 | Count | Mean Accuracy | Mean AUC | Mean Recall | Mean Precision |
| --- | --- | --- | --- | --- | --- | --- |
| no-ctr | ce | 16 | 0.8027 | **0.8477** | **0.7941** | 0.4548 |
| ctr-euc | ce | 16 | 0.8023 | 0.8329 | 0.7659 | 0.4532 |
| ctr-cos | ce | 16 | **0.8067** | 0.8427 | 0.7831 | **0.4599** |

On average, the use of supervised contrastive learning using cosine similarity (ctr-cos) techniques yielded the strongest result. We hypothesize that this is the case because using cosine similarity features for contrastive learning helps the model to maximize the angle between feature vectors of different classes and minimize the angle of feature vectors of the same class in multidimensional space, which is important for discerning between AD and CN.

A 2-dimensional PCA projection of the embedding dimension (16 features) for a random model is shown in **figure 3**. A visual analysis of these plots appear to show that the supervised contrastive learning models learned better embeddings with a more discriminative decision boundary for the AD and CN classes. This suggests that the model’s ability to generalize on a much larger data distribution will have improved as the location of the extracted features will be polarized and closer to the correct cluster corresponding to a specific class. The decision boundary will thus have an easier time differentiating between extracted features that belong to the AD and CN classes. Further analysis of model performance was done through the use of the Grad-CAM system [25] which allowed for the visual analysis of the important regions of the input scan which the model focused on. **Figure 4** shows that the model focused primarily on parts of the brain close to the hippocampal and prefrontal cortex regions - these regions of the brain are directly linked to memory and other cognitive abilities.

**Figure 3.**
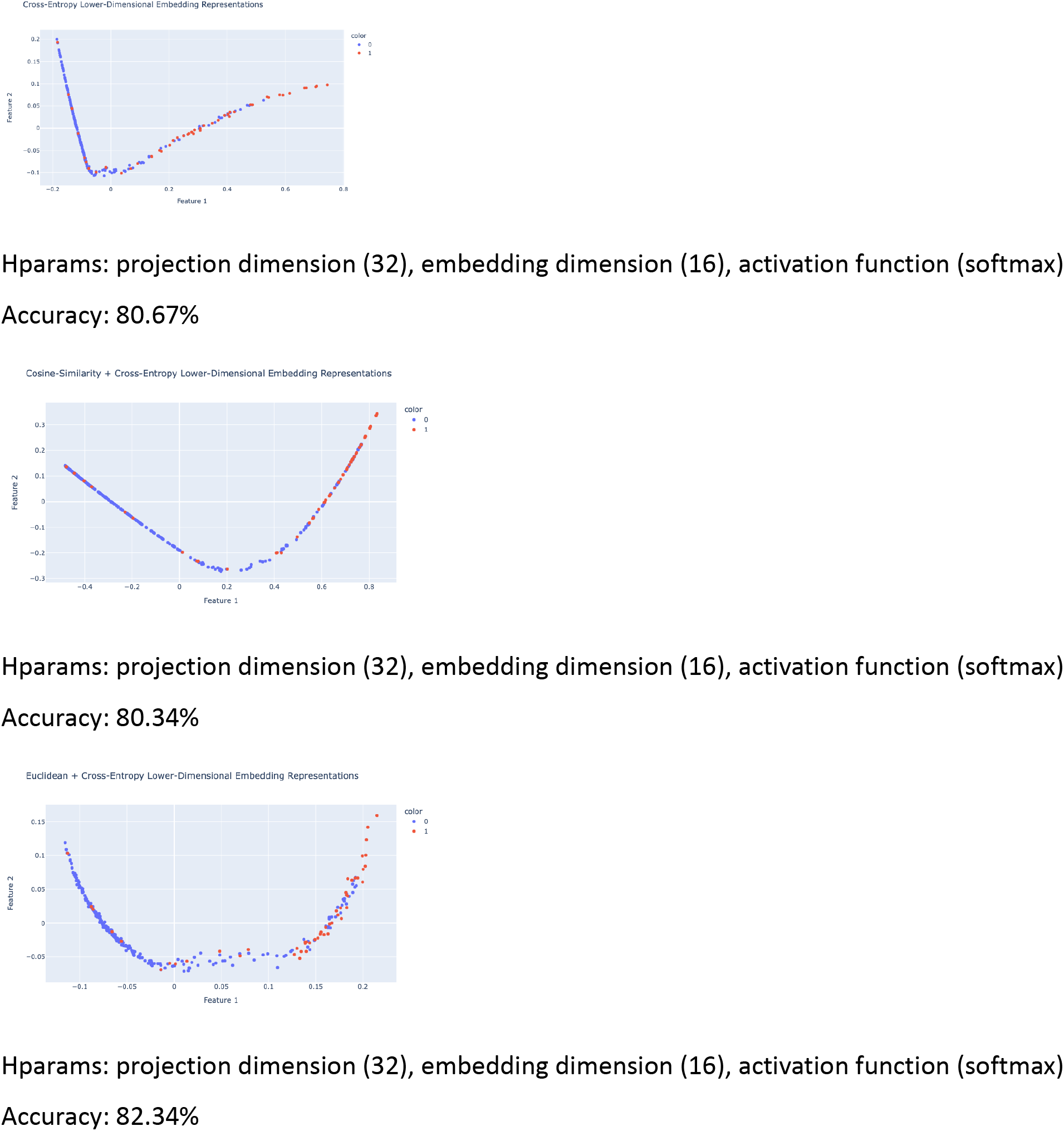
PCA Reduced Dimensionality Representation Analysis of Input Scans With and Without Contrastive Learning. To verify the functionality of the contrastive loss mechanism, a PCA visual analysis was performed on the extracted features of the test dataset distribution, where class 0 corresponds to CN and 1 corresponds to AD.

**Figure 4.**
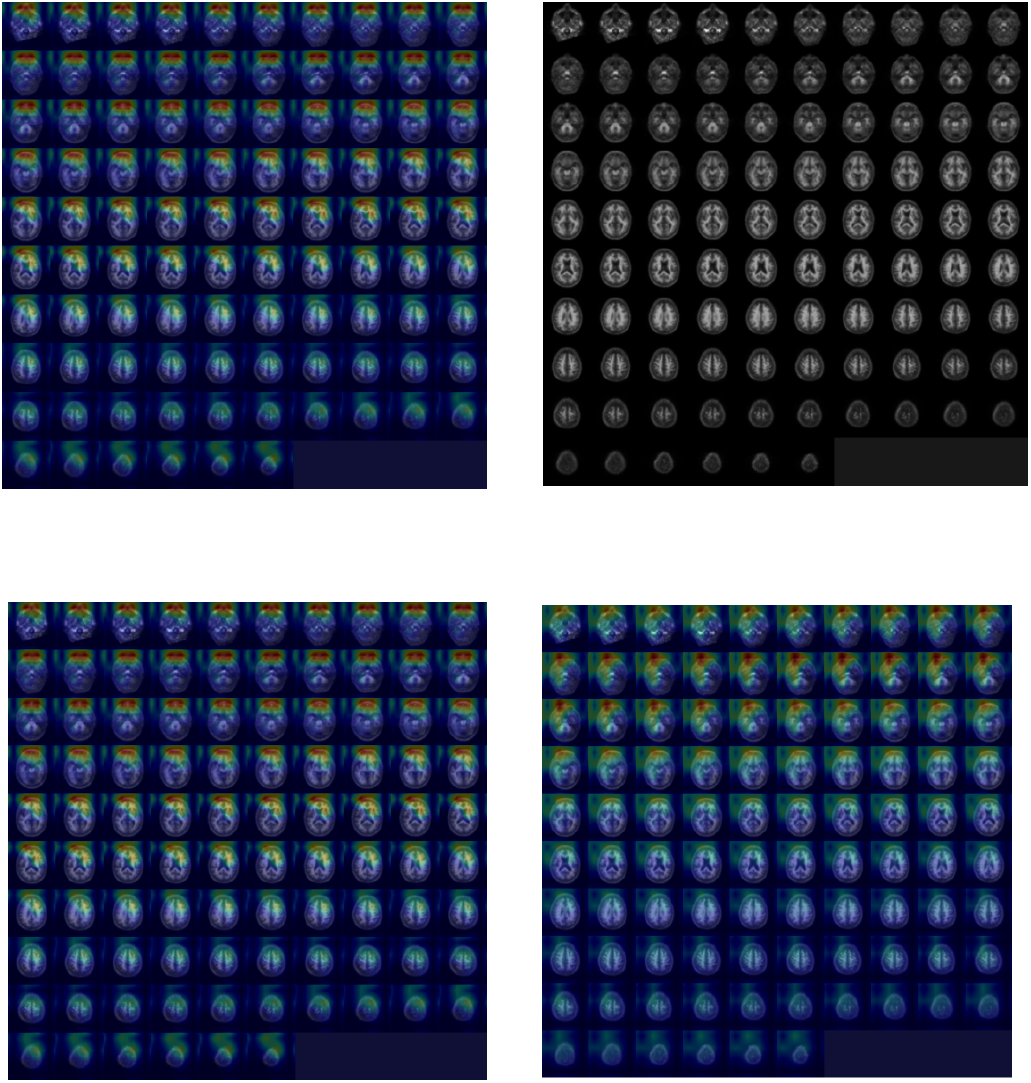
Grad-CAM Generated Image For Important Feature Visualization. Understanding whether the extracted features that the encoder portion of the classification system output actually contains useful features for the classification task is key to validate model performance. Grad-CAM is a technique that generates an interpretable gradient heatmap of what latent features are correlated to a given prediction. The figure above shows that the hippocampal region and prefrontal cortex is the region of the brain that the model most focuses on (red -> important; blue -> not important).

Overall, the results were highly informative with higher accuracies across mean, median, and max statistics for the models trained with supervised contrastive losses. Additionally, for our task, we found that the focal loss, batch size, and log loss were important hyperparameters for driving down the false negative rates and improving overall model performance.

## DISCUSSION

PET 18F-AV45 is an imaging modality frequently used within AD clinical diagnostics to allow radiologists to identify the correlations between beta-amyloid clusters in different regions of the brain, stemming from the hippocampal region. As AD progresses throughout its multiple stages, there is shown to be a positive correlation between the amount of secreted beta-amyloid plaque and the amount of time that has passed. Identifying relationships between beta-amyloid plaque amount, locations within the brain, and corresponding behavioral traits is a common diagnostic procedure used to effectively diagnose AD.

Contrastive learning optimizes the ability for a model to develop complicated decision boundaries for multi-class classification. Although not commonly present in diagnostic medicine literature, the results of this study show promise for improved DL-based diagnostics using contrastive learning, especially with constraints such as limited computational resources and small dataset sizes. The use of a contrastive learning system allowed the DL system to yield robust validation accuracies of 82%. The proposed tool has the potential to be deployed in clinical settings as a verification step to ensure that trained radiologist diagnoses are accurate. Additionally, a contrastive learning paradigm can be employed in other sectors of diagnostic medicine to automate the process or verify radiologist analyses.

Previous studies have utilized 3D CNNs for feature extraction and classification with the training process supported only by binary log loss. Cheng and Liu (2017) employed a dual modality strategy with MRI and PET to train an end-to-end 3D CNN + 2D CNN deep learning system. That study did not employ any other auxiliary loss and had a limited dataset size of 193 scans (93 AD and 100 CN). Similar studies accomplish automated dimensionality reduction to perform classification on latent space vectors through the use of autoencoder intermediaries to specialize in the feature extraction process. However, with a small dataset comes the issue of DL models having trouble generalizing across a larger data distribution during inference - something that needs to be held up to a high standard if the final goal is deployment in a clinical setting.

A thorough analysis of PCA plots showed that the positionings of extracted latent space vectors were optimized with clear visual distinctions between the regions of space occupied by the latent space vectors of the AD and the CN classes. Comparisons between the ground truth labels and predicted labels on the extracted latent space vectors plotted on multidimensional graphs show the model’s ability to effectively position vectors of varying classes and classify them accordingly. Through the use of an AE, the addition of a regularization term to prevent overfitting was no longer necessary as the reconstruction process acted as a regularizer for the classification head. To verify that the reconstruction mechanism was working properly, reconstructions results were visually analyzed using the image viewing environment on JupyterLab. Individual slices within the input and reconstructed output were extremely similar, an important observation considering the dimensionality of the input was reduced to [1, 32] and [1, 16] shaped latent feature vectors. The ability for the model to effectively reconstruct the input scan given a very small representation of the input scan supports the idea that the model was able to learn meaningful low-rank representations of the input.

In addition, the contrastive learning paradigm acted as another regularizer methodology alongside performing its primary task of optimizing latent space vector positions. Since the parameters of the AE feature extractor were being influenced by 3 different losses simultaneously, the issue of overfitting was not as substantial as shown in the validation results.

The limited amount of data available for medical imaging (PET and MRI modalities alike) has prevented proper benchmark comparisons to assess the statistical significance of a model’s performance in relation to that of another study’s model. Recreations of benchmark models are critical to ensure that all crucial variables are kept under control and a difference in dataset size or computational resources does not impact the model’s training process. Additionally, the PET 18F-AV45 scans used in this study were manually labeled by trained radiologists, a process that inherently contains subjective biases which could have resulted in inaccurate class prediction. Furthermore, hyperparameter tuning for weightage given to reconstruction, classification, and contrastive losses and other architecture design choices (e.g. the number of encoder/decoder blocks) will likely provide increased performances and stronger results.

## Data Availability

All data produced are available online at https://adni.loni.usc.edu/.

## Abbreviations

AD: Alzheimer’s Disease
CN: Control
PET 18F-AV45: florbetapir
ADNI: Alzheimer’s Disease Neuroimaging Initiative
DL: deep learning
CNN: convolutional neural network
AE: autoencoder
MSE: mean squared error
PCA: principal component analysis
ce: cross entropy
ctr: contrastive
euc: euclidean
cos: cosine similarity

